# Agile design and development of a high throughput cobas^®^ SARS-CoV-2 RT-PCR diagnostic test

**DOI:** 10.1101/2021.10.13.21264919

**Authors:** Chitra Manohar, Jingtao Sun, Peter Schlag, Chris Santini, Marcel Fontecha, Pirmin Lötscher, Carolin Bier, Kristina Goepfert, Dana Duncan, Gene Spier, Daniel Jarem, Dmitriy Kosarikov

**Affiliations:** Roche Molecular Systems, Inc., Pleasanton, United States; Roche Diagnostics International AG, Rotkreuz, Switzerland

**Keywords:** SARS-CoV-2, COVID-19, pandemic, RT-PCR, diagnostic testing, molecular assay design

## Abstract

Diagnostic testing is essential for management of the COVID-19 pandemic. An agile assay design methodology, optimized for the cobas^®^ 6800/8800 system, was used to develop a dual-target, qualitative SARS-CoV-2 RT-PCR test using commercially available reagents and existing sample processing and thermocycling profiles. The limit of detection was 0.004 to 0.007 TCID_50_/mL for USA-WA1/2020. Assay sensitivity was confirmed for SARS-CoV-2 variants Alpha, Beta, Gamma, Delta and Kappa. The coefficients of variation of the cycle threshold number (Ct) were between 1.1 and 2.2%. There was no difference in Ct using nasopharyngeal compared to oropharyngeal swabs in universal transport medium (UTM). A small increase in Ct was observed with specimens collected in cobas^®^ PCR medium compared to UTM. In silico analysis indicated that the dual-target test is capable of detecting all >1,800,000 SARS-CoV-2 sequences in the GISAID database. Our agile assay design approach facilitated rapid development and deployment of this SARS-CoV-2 RT-PCR test.

## 1. Introduction

A novel coronavirus, SARS-CoV-2, is the causative agent of COVID-19, a complex and potentially lethal human disease (1) that has infected over 200 million individuals worldwide as of August 2021 (2). The COVID-19 pandemic has been associated with more than four million deaths (2) and enormous economic impact across the world. SARS-CoV-2 is transmitted person-to-person via respiratory secretions, causing fever, respiratory symptoms (cough, shortness of breath), and subsequent immune system dysregulation. The clinical presentation of COVID-19 can vary from asymptomatic infection to mild illness to fatal disease (3-5).

The *Coronaviridae* is a family of viruses that cause illness ranging from mild respiratory infection (human coronaviruses 229E, NL63, HKU1, and OC43) to more severe diseases such as Middle East Respiratory Syndrome (MERS-CoV) and Severe Acute Respiratory Syndrome (SARS-CoV) (6). SARS-CoV-2 coronavirus belongs to the *Sarbecovirus* sub-genus, which also includes SARS-CoV and other betacoronaviridae identified in bats (7, 8).

Diagnostic testing is an essential component of infection prevention, control and disease management. One of the most sensitive types of diagnostic test currently available is based on specific detection of viral nucleic acids. One commonly used technology platform for such tests is real-time reverse transcription (RT)-PCR, which involves binding of primers and probes to specific regions of the pathogen’s genome. Rapid response to the need for testing for novel pathogens can be achieved by adaptation of existing automated instruments, well-established generic reagents and production facilities. The cobas^®^ 6800/8800 system (Roche Molecular Systems) is a widely used platform that supports the detection of many different clinically important viruses and bacteria using real-time PCR (9).

Early in the pandemic only a few SARS-CoV-2 genomic sequences were available in public databases (e.g. GISAID or NCBI). When the cobas^®^ SARS-CoV-2 test was designed in early 2020, little was known about what regions of the genome might be subject to sequence variation and/or recombination. Despite the paucity of knowledge about potential sequence variation, we designed a single well, dual-target assay to detect SARS-CoV-2-specific sequences using targets in the non-structural region of the ORF1a/b locus, and a conserved region in the structural envelope (E)-gene common to all sarbecoviruses, including SARS-CoV-2. The test was designed to meet the need for high throughput testing on the cobas 6800/8800 system, which performs fully automated sample preparation, real time RT-PCR reaction setup, target amplification and detection.

SARS-CoV-2 can evolve in response to external selection pressures. Strong but incomplete inhibition of replication, which might occur in an infected person with partial immunity, can result in the selection of SARS-CoV-2 variants that have higher replicative fitness than the wild-type virus in a population of susceptible hosts. Similarly, if a naturally occurring variant were to arise with increased ability to spread in an immunologically naïve population, it could out-compete the wild-type virus in a relatively short period of time. The emergence of several “variants of concern” (VOC) and “variants of interest” in many different locations of the world in recent months is therefore not unexpected, and has several important public health and clinical implications (10-16). Sequence variation in such variants has the potential to interfere with molecular diagnostic test performance.

## 2. Methods

### 2.1 Viruses

An isolate of SARS-CoV-2 from the first patient diagnosed with COVID-19 in the US (USA-WA1/2020, catalog number NR-52281, lot number 70033175, 2.8×10^5^ TCID_50_/mL) (17) was obtained from BEI Resources (Manassas, VA). Based on information provided in the Certificate of Analysis from the vendor, one TCID_50_/mL is equal to 7393 genome equivalents (RNA copies) by droplet digital PCR™ (Bio-Rad^®^). An isolate of SARS-CoV-2 from a German patient (BetaCoV/Germany/BavPat1/2020, catalog number 026V-03883, 3.2 × 10^6^ pfu/mL) was obtained from the European Virus Archive Global (Marseille, France). All experiments with replication competent SARS-CoV-2 were performed in a biocontainment level 3 facility in Switzerland. Virus stocks were diluted in a simulated matrix, consisting of human cells and mucin in Universal Viral Transport Medium (UTM, Copan Diagnostics, Murrieta, CA; https://www.copanusa.com/wp-content/uploads/2020/02/UTM-Package-Insert.pdf), that was shown to be equivalent to natural nasopharyngeal matrix in assay performance (data not shown). Virus stocks for variants of concern were obtained from BEI Resources (catalog numbers NR-54000, NR-54008, NR-54982, NR-55611, NR-55486, NR-55308, NR-55309, NR-55439, NR-55469, and NR-55654). For the wild-type strain, genomic RNA was used (NR-52499). Virus RNA concentration was determined by droplet digital PCR.

### 2.2 cobas SARS-CoV-2 assay design

The cobas 6800/8800 platform is an end-to-end system that includes hardware, software, reagents and consumables that performs automated nucleic acid testing. The platform is intended for moderate-to high-throughput laboratories, where a large number of test results are needed within short periods of time. Different tests that are performed on the cobas 6800/8800 system use the same generic reagents and share common sample processing and PCR profiles coupled with target-specific assay oligonucleotides and positive control. Therefore, cobas SARS-CoV-2 was developed using the automated, well established conditions for the cobas 6800/8800 system, sample preparation workflow and thermal cycling profile, allowing for simultaneous inclusion of other diagnostic tests designed for the cobas 6800/8800 platform (amplification/detection on the same PCR plate). All reagents were developed using synergies whenever possible, such as common raw material, manufacturing and use test (kit release) procedures, controls and PCR reaction master mix formulation.

To design primers and probes for the PCR assay, Agile Assay Design (AAD) software (Roche Molecular Systems) was used to select optimal oligonucleotide length and sequence based on the seven available SARS-CoV-2 sequences in the Global Initiative on Sharing All Influenza Data (GISAID, www.gisaid.org) (18). The designs took into consideration key PCR parameters that predict efficient assay performance using Roche master mix and reagents on the cobas 6800/8800 system. To evaluate inclusivity, the delay in cycle threshold (dCt) compared to perfectly matching primers was modeled. The algorithm was based on experiments performed with 20 perfectly matched primers at different locations and 268 corresponding, mis-matched primers containing one to six nucleotide mismatches, using both DNA and RNA templates to experimentally measure dCt. The models use thermodynamics parameters (free energies, dG) for DNA:DNA interactions (19). The melting temperatures for probes were calculated using Melting5 software (20) at 100 nM probe concentration, 1.7 mM Mg^2+^ and 50mM Na^+^/K^+^.

### 2.3 Inclusivity

Inclusivity analysis was performed using all available SARS-CoV-2 sequences in GISAID as of June 15, 2021 (n= 1,874,933). The predicted impact of each variant Target 1 and Target 2 primer and probe binding site sequence (six sites) was evaluated using the AAD software and quantitated as the predicted increase in Ct or probe melting temperature (Tm).

### 2.4 Sensitivity (Limit of Detection)

To determine the limit of detection (LoD), USA-WA1/2020 live virus was serially diluted in simulated clinical matrix. A total of seven concentrations, generated using 3-fold serial dilutions of the stock virus, were tested, with a total of 21 replicates per concentration and an additional 10 replicates of diluent only. LoD was determined by probit analysis based on the titer given by the supplier and dilution factor. The LoD was confirmed using a second virus isolate, BetaCoV/Munich/BavPat1/2020, similarly serially diluted.

### 2.5 Precision

Precision was assessed with a panel made using cultured SARS-CoV-2 (USA-WA1/2020, heat-inactivated) in simulated clinical matrix in UTM. SARS-CoV-2 virus stock material was serially diluted to generate a panel consisting of three concentration levels (weak, low and moderate positive) corresponding to approximately 0.3x, 1x and 3x the LoD, respectively. The samples were tested over 15 days, three reagent lots, on three instruments and by three operators. Each test day, two runs were performed per lot and per instrument, using three replicates per panel member per run. A total of 90 replicates per concentration level were tested over the course of the study. The 90 replicates were distributed across three reagent lots (30 replicates each) and three cobas^®^ 6800/8800 Systems.

### 2.6 Matrix/Collection media equivalency

The relative performance of different specimen types or transport media was evaluated using cultured virus (USA-WA1/2020 strain), spiked into paired specimens from SARS-CoV-2 negative individuals with symptoms of an upper respiratory infection collected in 2018 (before the SARS-CoV-2 outbreak) and stored frozen at -80°C. The final virus concentration was approximately 0.054 TCID_50_/mL, or 1.5 times higher than the LoD. The relative performance of nasopharyngeal swab (NPS) and oropharyngeal swab (OPS) specimens was compared in universal viral transport medium (BD, Franklin Lakes, NJ; equivalent to UTM). NPS swabs were used for the comparison between UTM or the virus-inactivating cobas^®^ PCR Media (CPM; Roche Molecular Systems) (21). A total of 21 replicates at 0.054 TCID_50_/mL were tested. Similarly, detection of SARS-CoV-2 RNA in specimens collected using polyester woven or nylon flocked swabs in UTM or 0.9% saline physiological solution was evaluated using cultured virus (USA-WA1/2020 strain) spiked into matched nasal swab (NS) specimens from SARS-CoV-2 negative individuals. Three specimens were self-collected from each of 45 healthy donors: two using either a woven polyester or nylon flocked swab and placed in UTM, and one (from the other nostril) using a woven polyester swab placed in 0.9% physiological saline. A total of 17 replicates at 0.054 TCID_50_/mL were tested.

## 3. Results

### 3.1 Assay Design

At the time when the cobas SARS-CoV-2 test was designed (January 2020), only seven SARS-CoV-2 sequences were available from the Global Initiative on Sharing All Influenza Data (GISAID). To guard against the possibility that viral sequence changes in the target site may negatively impact detection, two complementary strategies were employed. First, a dual target design was chosen, which dramatically increases the likelihood of viral sequence detection and PCR signal preservation from one of the two targets even if the other has sequence variations. Second, selection of target regions where sequences are conserved between virus species (e.g. between SARS-CoV-2 and SARS-CoV) greatly enhances the chance that this sequence will remain conserved within species. We selected one target (Target 1) in the ORF1a/b coding region that is highly specific for SARS-CoV-2. The second target (Target 2) is located in a region of the E gene that is conserved among sarbecoviruses (Fig. 1). A proprietary software program, AAD (see Methods), was used to evaluate the seven available SARS-CoV-2 sequences and over 1000 sequences from other sarbecoviruses, including SARS-CoV, that were available from NCBI. Target 2 positivity can be interpreted unambiguously given the clinical and epidemiological context (i.e. with knowledge that SARS-CoV-2 is circulating but SARS-CoV is not).

**Figure 1.**
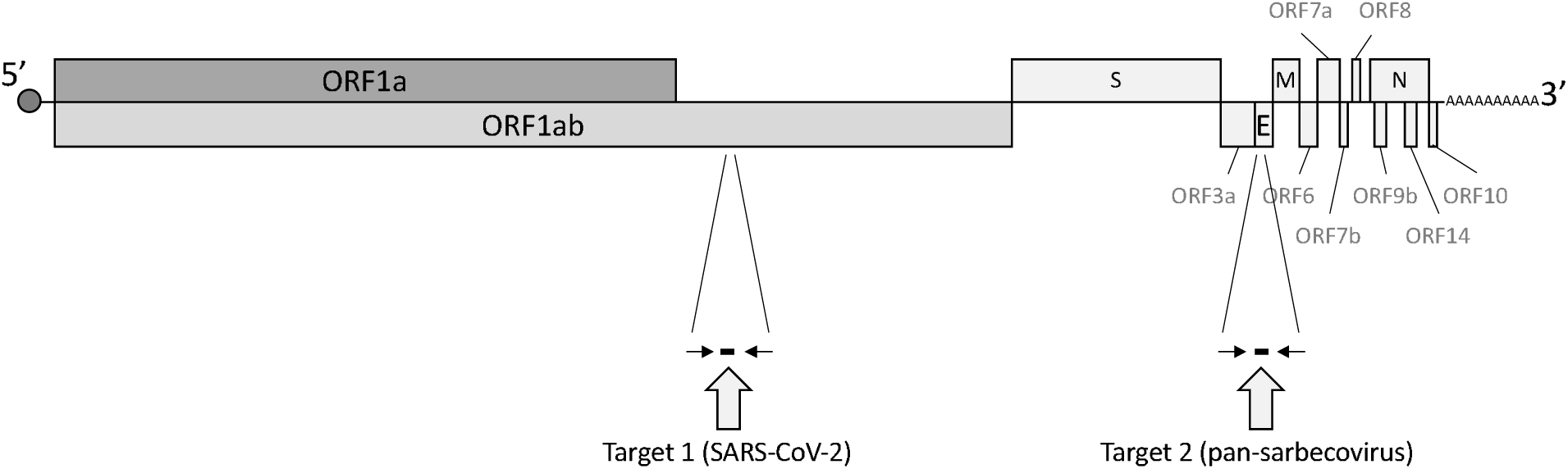
SARS-CoV-2 genome diagram and location of cobas assay target regions. ORF: open reading frame; S: spike; E: envelope; M: matrix; N: nucleocapsid. Forward and reverse primers are represented by arrows, and probes by black rectangles, for Target 1 (ORF1a/b) and 2 (E gene).

### 3.2 Inclusivity

In June 2021, an updated *in silico* analysis of 1,874,933 SARS-CoV-2 sequences in the GISAID database was performed. Table 1 summarizes the predicted impact of sequence variation at primer or probe binding sites for variants represented in at least 0.02% (403 or more) of all SARS-CoV-2 sequences in the GISAID database (a complete listing of all variants including very infrequent sequences can be found in the Supplemental Data Table S1). A summary of the numbers of individual haplotypes (defined as a specific sequence including the four primer and two probe binding sites) is shown in Table 2. Overall, 98.56% of sequences have no changes in primer or probe binding sites at either target, and 1.4% have only a single change. One of these (variant 13, found in 0.025% of sequences), which has a single change in the probe binding site for Target 2 (Table 1), is predicted to reduce the reactivity of single probes or primers, and has been previously shown to be associated with failure of detection at Target 2 (22). An additional 74 (0.004%) sequences, bearing between two and eight nucleotide changes, were observed that are predicted to impact assay performance for target 1 (n=31) or target 2 (n=43; Table 2). No variant reported in the GISAID database had changes in both target regions simultaneously. Therefore no impact on cobas SARS-CoV-2 test performance is anticipated, considering all the sequences available.

**Table 1.**
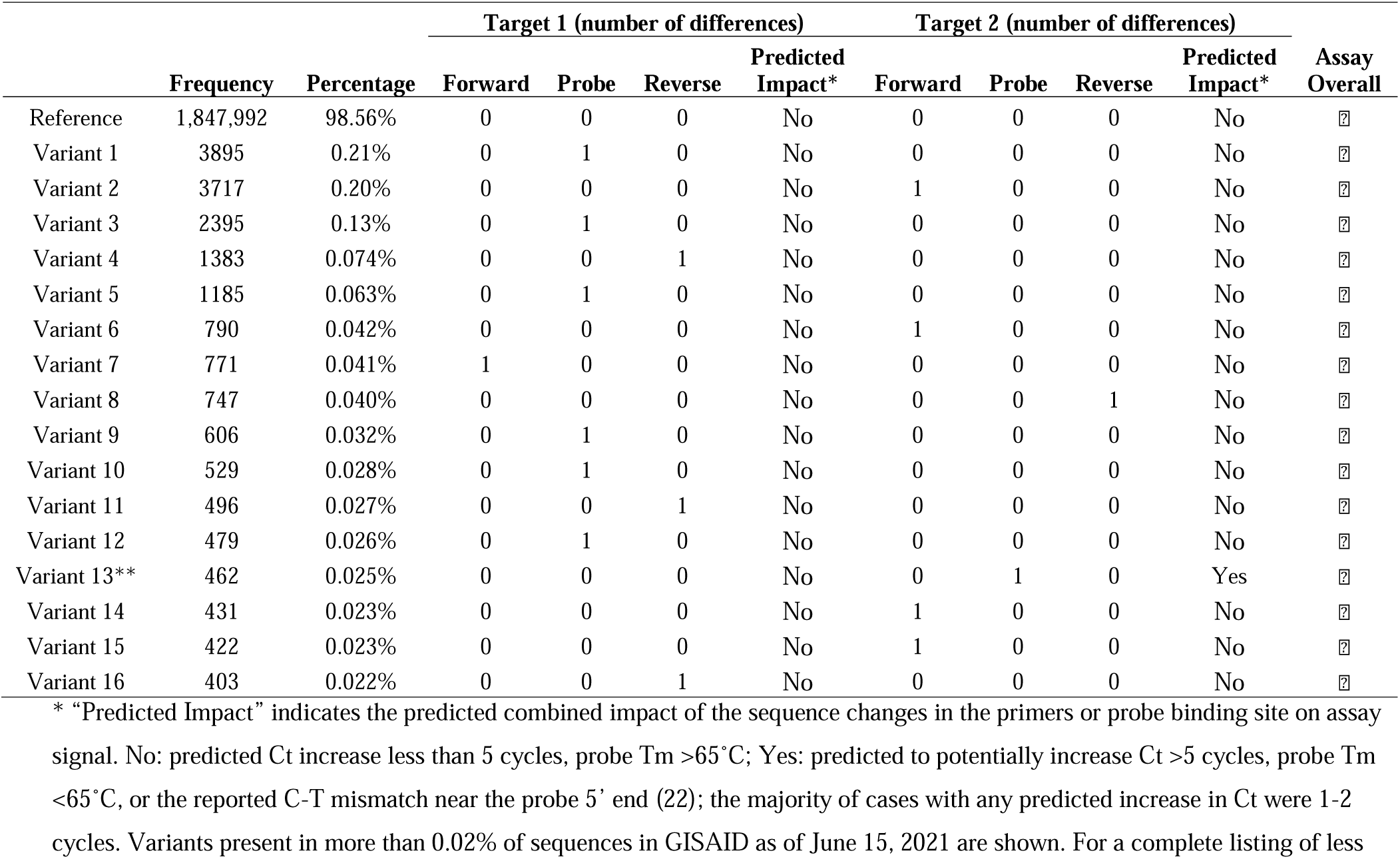
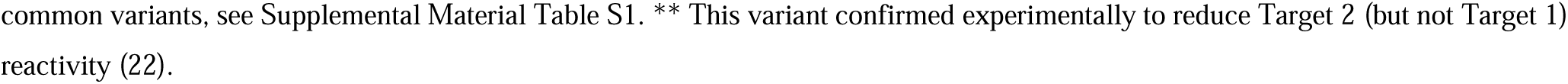
Analysis of cobas SARS-CoV-2 primer and probe binding site sequence variation.

**Table 2.**
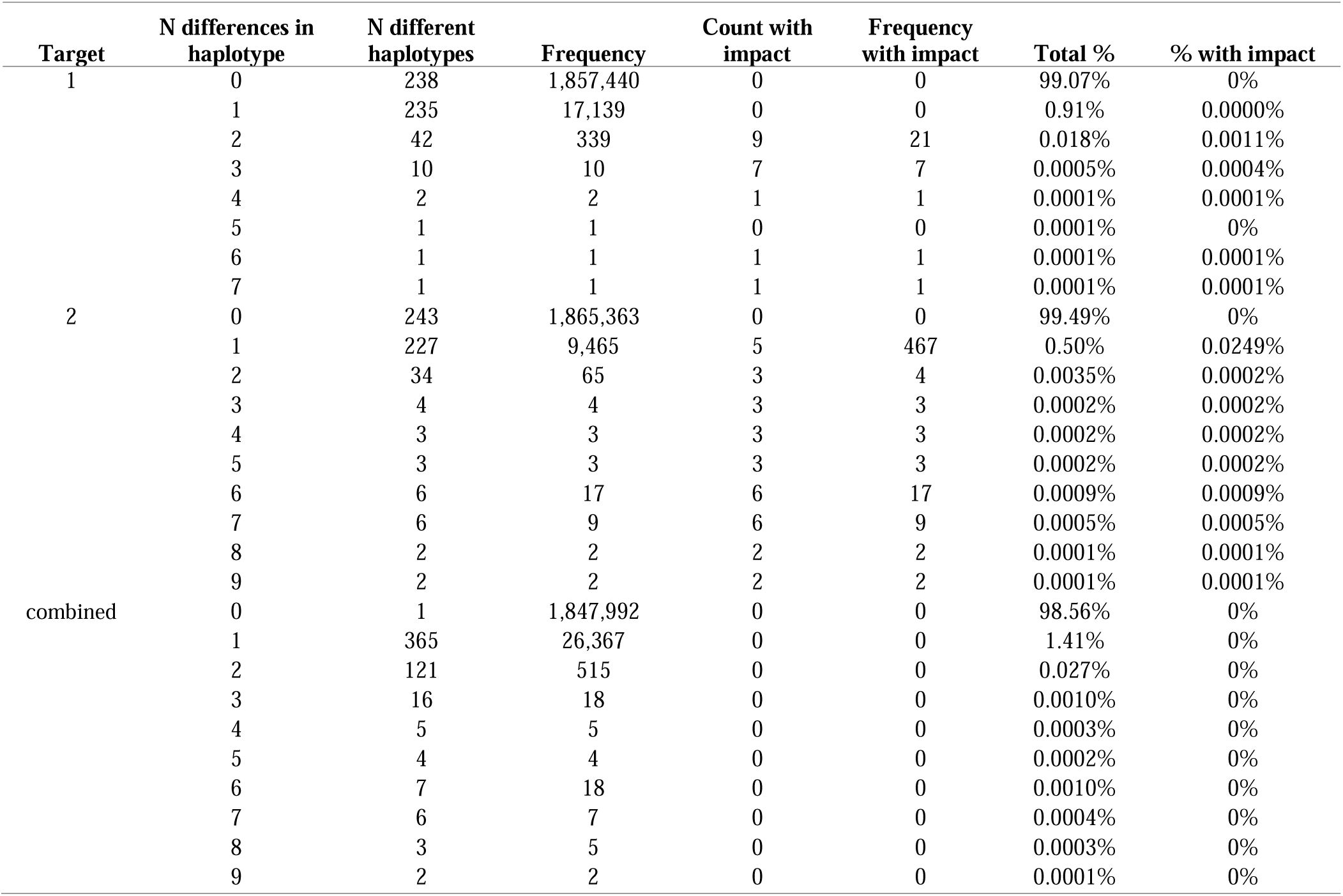
Summary of Assay Inclusivity.

### 3.3 Sensitivity (Limit of Detection)

Assay sensitivity was determined by replicate testing of serial dilutions of USA-WA1/2020 virus. As shown in Table 3, the concentration level with observed test positivity rates ≥95% were 0.009 and 0.003 TCID_50_/mL for Target 1 and 2, respectively. The probit model predicted 95% test positivity rates at virus titers of 0.007 (95% CI: 0.005 – 0.036) and 0.004 (95% CI: 0.002 – 0.009) TCID_50_/mL for Target 1 and 2, respectively. Based on an estimate of the number of RNA copies per TCID_50_ in the virus stock used for this experiment, this corresponds to 52 copies/mL for Target 1 and 30 copies/mL for Target 2.

**Table 3.** Limit of detection

| Concentration<br>(TCID<sub>50</sub>/mL) | RNA<br>concentration<br>(copies/mL)* | Total valid<br>results | Test positivity (%)† |  | Mean Ct‡ |  |
| --- | --- | --- | --- | --- | --- | --- |
|  |  |  | Target 1 | Target 2 | Target 1 | Target 2 |
| 0.084 | 621 | 21 | 100 | 100 | 31.0 | 33.0 |
| 0.028 | 207 | 21 | 100 | 100 | 31.8 | 34.1 |
| 0.009 | 67 | 21 | 100 | 100 | 32.7 | 35.2 |
| 0.003 | 22 | 21 | 38.1 | 100 | 33.5 | 36.4 |
| 0.001 | 7.4 | 21 | 0 | 52.4 | n/a | 37.9 |
| 0.0003 | 2.2 | 21 | 0 | 14.3 | n/a | 37.2 |
| 0.0001 | 0.7 | 21 | 0 | 9.5 | n/a | 38.5 |
| 0 (blank) | 0 | 10 | 0 | 0 | n/a | n/a |

\* Conversion from TCID<sub>50</sub>/mL to RNA copies/mL based on information provided in the Certificate of Analysis from the vendor: one
TCID<sub>50</sub>/mL is equal to 7393 genome equivalents (RNA copies) by droplet digital PCR
† All replicates where Target 1 was positive were also positive for Target 2.
‡ Calculations only include positive results.

A second sensitivity study was performed using a different virus isolate (BetaCoV/Germany/BavPat1/2020) whose concentration is reported in plaque forming units (pfu) instead of TCID_50_. The concentration levels with observed test positivity rates greater than or equal to 95% using this isolate were 0.011 pfu/mL for Target 1 and 0.004 pfu/mL for Target 2 (data not shown). Probit analysis predicted 95% test positivity rates at virus titers of 0.007 pfu/mL (95% CI: 0.005 – 0.023) for Target 1 and 0.004 pfu/mL (95% CI: 0.002 – 0.009) for Target 2.

Assay sensitivity for SARS-CoV-2 variants of concern alpha, beta, gamma, and delta (B.1.1.7, B.1.351, P.1, and B.1.617.2, respectively), variant of interest lambda (C.37), and several variants under monitoring (some of which were formerly variants of interest) was confirmed by testing 5 to 8 replicates of three different dilutions of virus stocks for each variant near the LoD determined above (50 to 250 copies/mL). For Target 1, at the two highest concentrations, all results were positive for all variants (Table 4). At 50 copies/mL for Target 1, 62.5% (beta), 87.5% (wild-type, alpha and gamma) or 100% (delta, kappa) of results were positive. At all concentrations tested, all results were positive for Target 2.

**Table 4.**
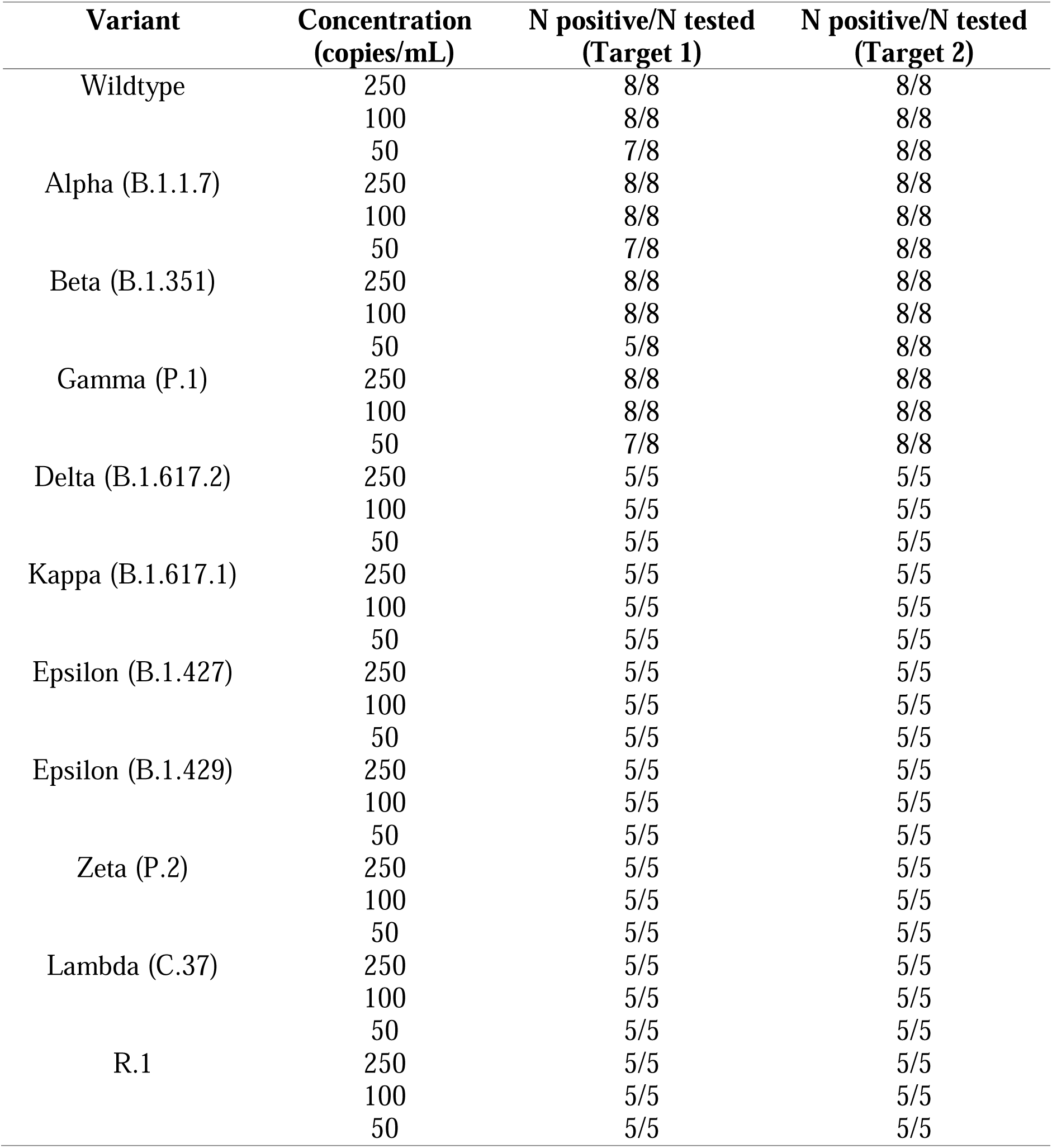
Detection of variants of concern, variants of interest and variants under monitoring

| **Variant** | **Concentration**<br>**(copies/mL)** | **N positive/N tested**<br>**(Target 1)** | **N positive/N tested**<br>**(Target 2)** |
| --- | --- | --- | --- |
| Wildtype | 250 | 8/8 | 8/8 |
|  | 100 | 8/8 | 8/8 |
|  | 50 | 7/8 | 8/8 |
| Alpha (B.1.1.7) | 250 | 8/8 | 8/8 |
|  | 100 | 8/8 | 8/8 |
|  | 50 | 7/8 | 8/8 |
| Beta (B.1.351) | 250 | 8/8 | 8/8 |
|  | 100 | 8/8 | 8/8 |
|  | 50 | 5/8 | 8/8 |
| Gamma (P.1) | 250 | 8/8 | 8/8 |
|  | 100 | 8/8 | 8/8 |
|  | 50 | 7/8 | 8/8 |
| Delta (B.1.617.2) | 250 | 5/5 | 5/5 |
|  | 100 | 5/5 | 5/5 |
|  | 50 | 5/5 | 5/5 |
| Kappa (B.1.617.1) | 250 | 5/5 | 5/5 |
|  | 100 | 5/5 | 5/5 |
|  | 50 | 5/5 | 5/5 |
| Epsilon (B.1.427) | 250 | 5/5 | 5/5 |
|  | 100 | 5/5 | 5/5 |
|  | 50 | 5/5 | 5/5 |
| Epsilon (B.1.429) | 250 | 5/5 | 5/5 |
|  | 100 | 5/5 | 5/5 |
|  | 50 | 5/5 | 5/5 |
| Zeta (P.2) | 250 | 5/5 | 5/5 |
|  | 100 | 5/5 | 5/5 |
|  | 50 | 5/5 | 5/5 |
| Lambda (C.37) | 250 | 5/5 | 5/5 |
|  | 100 | 5/5 | 5/5 |
|  | 50 | 5/5 | 5/5 |
| R.1 | 250 | 5/5 | 5/5 |
|  | 100 | 5/5 | 5/5 |
|  | 50 | 5/5 | 5/5 |

### 3.4 Precision

Summary statistics for Ct values for the weak (∼0.3x), low (∼1.0x), and moderate (∼3.0x) positive concentration levels by variance component are shown in Table 5. Coefficients of variation of less than 2.0% CV were observed for all variables and concentration levels. Slightly more variability was observed between reagent lots and in the within-run residual category. Precision values of 0.8% CV or less were observed between instruments, day-to-day, and run-to-run. Overall, the coefficients of variation ranged from 1.1 to 1.9% for Target 1, and from 1.1 to 2.2% for Target 2.

**Table 5.** Assay precision

| Target | Level (x<br>LoD) | Hit rate | Mean Ct | Instrument<br>to-Instrument |  | Lot-to-Lot |  | Day-to-Day |  | Run-to-Run |  | Within-Run<br>(Residual) |  | Total |  |
| --- | --- | --- | --- | --- | --- | --- | --- | --- | --- | --- | --- | --- | --- | --- | --- |
|  |  |  |  | SD | CV% | SD | CV% | SD | CV% | SD | CV% | SD | CV% | SD | CV% |
| Target 1 | ~0.3x | 10.0% | 32.5 | 0.0 | 0.0 | 0.0 | 0.0 | 0.0 | 0.0 | 0.0 | 0.0 | 0.5 | 1.4 | 0.5 | 1.4 |
|  | ~1.0x | 91.1% | 32.1 | 0.0 | 0.0 | 0.2 | 0.6 | 0.1 | 0.3 | 0.0 | 0.0 | 0.6 | 1.8 | 0.6 | 1.9 |
|  | ~3.0x | 100.0% | 31.2 | 0.0 | 0.0 | 0.2 | 0.7 | 0.0 | 0.0 | 0.0 | 0.0 | 0.3 | 0.9 | 0.4 | 1.1 |
| Target 2 | ~0.3x | 34.4% | 35.4 | 0.0 | 0.0 | 0.5 | 1.3 | 0.3 | 0.8 | 0.1 | 0.2 | 0.5 | 1.5 | 0.8 | 2.2 |
|  | ~1.0x | 93.3% | 34.2 | 0.0 | 0.0 | 0.1 | 0.3 | 0.2 | 0.6 | 0.0 | 0.0 | 0.7 | 2.0 | 0.7 | 2.2 |
|  | ~3.0x | 100.0% | 32.9 | 0.0 | 0.1 | 0.0 | 0.0 | 0.0 | 0.0 | 0.0 | 0.0 | 0.4 | 1.1 | 0.4 | 1.1 |

### 3.5 Matrix/Collection media equivalency

Nasopharyngeal swab (NPS) and oropharyngeal swab (OPS) specimens are suitable for use in the diagnosis of respiratory virus infections. To demonstrate matrix equivalency, cultured virus (USA-WA1/2020 strain) was spiked into paired OPS or NPS specimens from SARS-CoV-2 negative individuals to final concentrations of approximately 0.054 TCID_50_/mL, or 1.5 times higher than the LoD. The cycle threshold number (Ct, inversely correlated with RNA quantity in the specimen) from replicate tests for each target is shown in Fig. 2. There was no statistically significant difference between mean Ct values in NPS vs OPS for either target (paired t-test *P* value >0.1).

**Figure 2.**
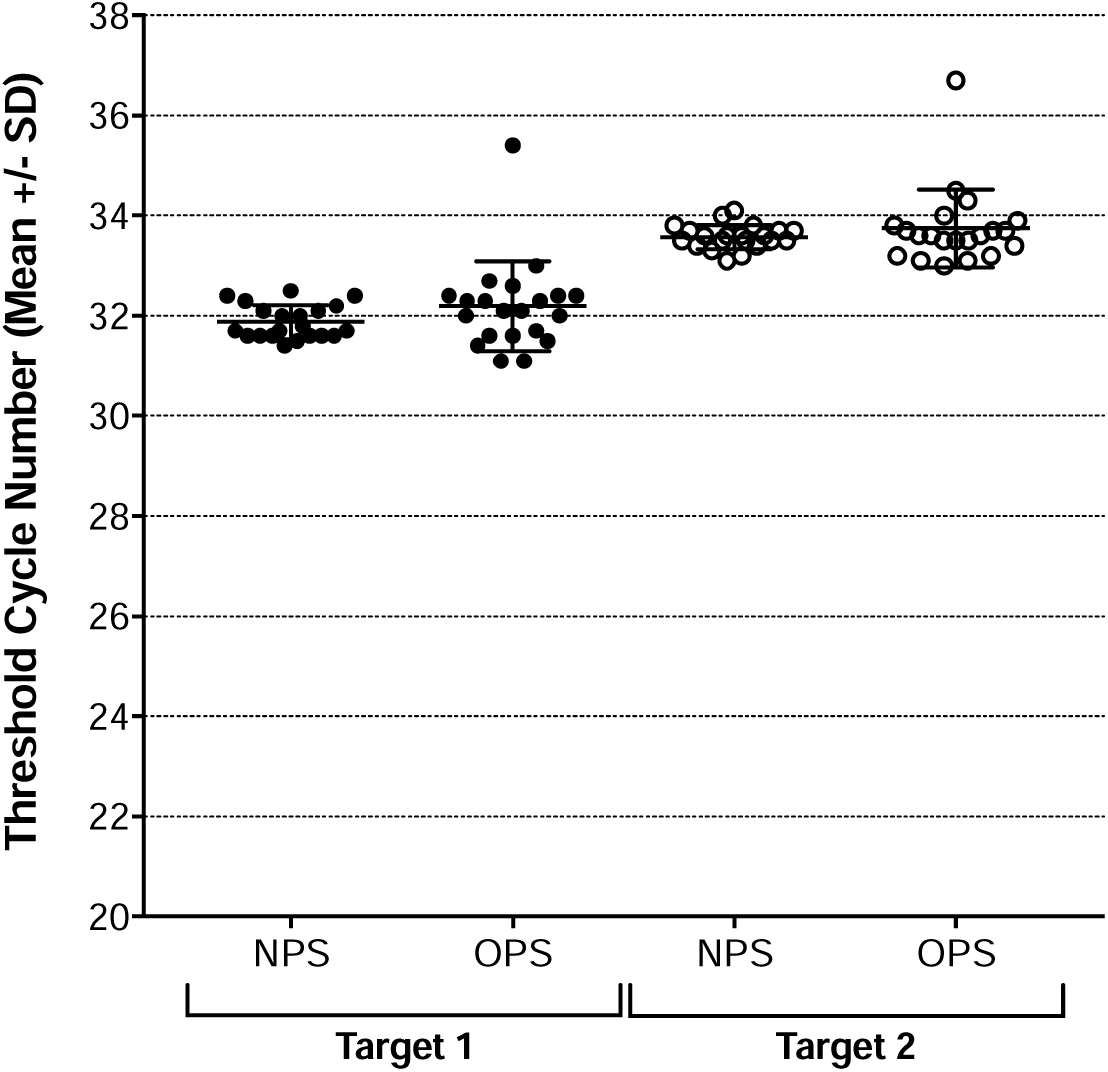
Specimen type equivalency. Individual results are plotted for each specimen type and target; horizontal bars represent the mean and standard deviation (SD) for each group. NPS: nasopharyngeal swab; OPS: oropharyngeal swab. Target 1 (filled circles) and Target 2 (open circles) are shown separately.

Similar experiments were performed to compare the Ct values in specimens collected in different types of swabs and collection media. Ct values for specimens diluted in CPM were slightly higher (difference in mean Ct of 0.4 to 0.7) than in Universal Transport Media (UTM) for both targets (paired t-test *P* value ≤0.002; Fig. 3A). Specimens collected using nylon flocked swabs in UTM yielded a minimal increase in mean Ct vs. polyester woven swabs in the same medium (difference 0.6 Ct, paired t-test *P* value 0.0065), while polyester woven swabs in saline yielded similar Ct values compared to the same type of swab in UTM (paired t-test *P* value >0.4; Fig. 3B).

**Figure 3.**
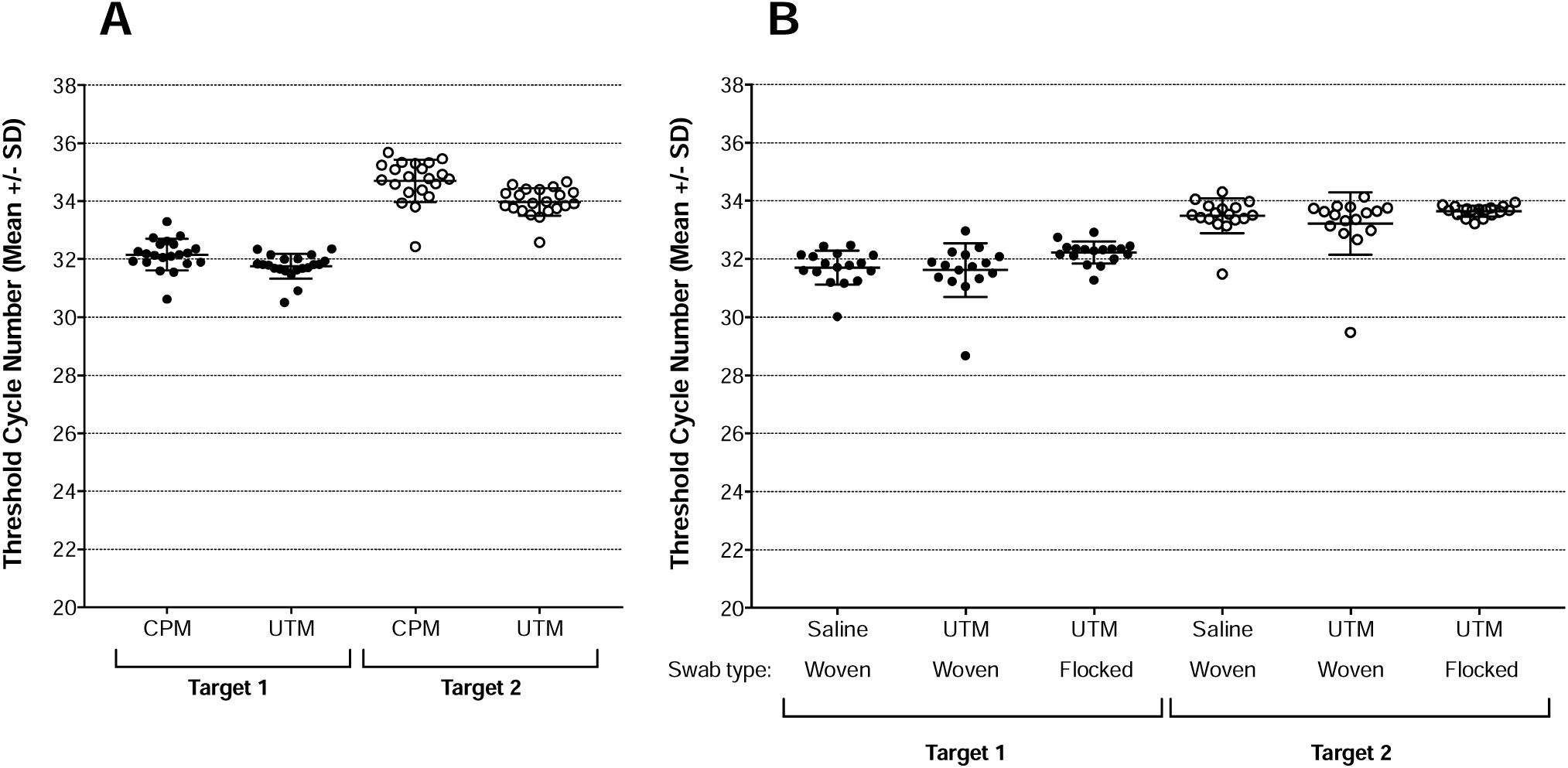
Specimen collection matrix equivalency. **A**: CPM vs. UTM. **B**. flocked or woven swabs in UTM and woven swabs in saline. Individual results are plotted for each matrix and target; horizontal bars represent the mean and standard deviation (SD) for each group. CPM: cobas PCR medium; UTM: universal transport medium. Target 1 (filled circles) and Target 2 (open circles) are shown separately.

## 4. Discussion

Deployment of new diagnostic tests for emerging pathogens in a timely manner is an integral part of the public health response to pandemics, such as the one caused by infection with SARS-CoV-2. Several different factors can be leveraged to speed development and deployment of novel diagnostic assays at the start of a pandemic or any emerging infectious disease. When tests for new targets can be deployed using existing infrastructure central testing laboratories can capitalize on already deployed instruments and trained personnel. The cobas 6800/8800 platform represents such an opportunity, since real-time PCR primers and probes can be designed to work in the context of existing, well-characterized assay chemistry and conditions. The AAD software for primer/probe design is an effective tool that facilitates rapid assay development for new pathogens. In addition, when reference materials are available for preparation of contrived specimens in a variety of authentic and simulated clinical matrices, initial performance evaluations required for emergency use authorization can be completed quickly. The convergence of these features enabled the rapid development of this diagnostic test, which was the first such test granted emergency use authorization in the US in early 2020. The paucity of sequence data at the early stages of an outbreak is a significant challenge for molecular test development (23, 24). The cobas SARS-CoV-2 test primers and probes were designed at a time when only seven genomic sequences were publicly available. However, sequence conservation may be predicted by considering data from related viruses for which more sequences have been characterized. A region of conserved sequence across different virus species is also likely to remain conserved within a species in the future.

Our approach included two different sets of primers and probes, one of which (Target 1) is specific for ORF1a/1b of SARS-CoV-2, while the other (Target 2) is intended to react with E-gene sequences of SARS-CoV-2, SARS-CoV, and other sarbecoviruses that infect bats. The genes in which the targets should be located were not pre-determined, but instead the AAD approach identified sites anywhere in the genome predicted to provide the best performance and desired level of sequence conservation. While Target 2 reactivity is expected for both SARS-CoV and SARS-CoV-2, virus positivity can be unambiguously established with knowledge of extant virus and disease prevalence, and in combination with the SARS-CoV-2 specific Target 1 result. Importantly, the use of two targets enables test accuracy even in the presence of sequence variation in one of the two target sequences. This has been demonstrated to occur in at least one case (22).

The limit of detection of the cobas SARS-CoV-2 test was determined to be between 0.004 and TCID_50_/mL or between 30 and 52 copies/mL for the particular virus stock used here. It should be noted that the relationship between TCID_50_ (or plaque forming units) and number of RNA copies may differ between virus preparations; this may explain small differences in LoD reported in RNA copies/mL elsewhere (25). A lower LoD for the E-gene target compared to other targets has been reported previously (26), which is consistent with our results. We noted that at low virus input levels, Target 1 positivity is impacted more than Target 2, in spite of higher Ct values for Target 2.

Previous evaluations have provided conflicting results regarding the relative sensitivity of SARS-CoV-2 RT-PCR assays using NPS, OPS or other specimen types (27-34). Our results support the use of either NPS or OPS as the specimen type, since there was no difference in the ability to detect SARS-CoV-2 RNA spiked into either specimen at low levels. While NPS is viewed as the gold standard for many respiratory pathogens, OPS are easier to obtain and less intrusive for the patient. These findings should be confirmed with specimens from infected individuals in the clinic.

In the early phases of the COVID-19 pandemic, many clinicians experienced a shortage of recommended sample collection materials including media for specimen storage and shipping. Our data indicate equivalent sensitivity of the cobas SARS-CoV-2 test when specimens are stored in UTM, CPM or saline, as long as specimens are refrigerated (2-8 °C) and stored for 6 days or less. CPM has the added advantage of inactivating the infectivity of SARS-CoV-2, thus improving biosafety for specimen handling in the laboratory (21).

Several independent studies have reported on the performance of the cobas SARS-CoV-2 test, in comparison with laboratory-developed tests (LDTs) or other commercial assays. Generally, overall percent agreement values range from 95 to 99% (25, 26, 35-37), with more discordance observed in specimens with low viral loads (35, 36, 38).

In conclusion, the cobas SARS-CoV-2 is a robust, sensitive and specific test for qualitative diagnosis of infection by SARS-CoV-2 that can be performed using equipment and infrastructure already widely available in clinical reference laboratories globally. The rapid development and deployment of this test was made possible by the application of the AAD approach and early availability of sequence information and reference reagents.

## Data Availability

All data produced in the present study are available upon reasonable request to the authors

## Acknowledgements

This publication was supported by the European Virus Archive GLOBAL (EVA-GLOBAL) project that has received funding from the European Union’s Horizon 2020 research and innovation programme under grant agreement No 871029. Manuscript preparation services were provided by Data First Consulting (Sebastopol, CA).

